# Rapid quantitative screening assay for SARS-CoV-2 neutralizing antibodies using HiBiT-tagged virus-like particles

**DOI:** 10.1101/2020.07.20.20158410

**Authors:** Kei Miyakawa, Sundararaj Stanleyraj Jeremiah, Norihisa Ohtake, Satoko Matsunaga, Yutaro Yamaoka, Mayuko Nishi, Takeshi Morita, Ryo Saji, Mototsugu Nishii, Hirokazu Kimura, Hideki Hasegawa, Ichiro Takeuchi, Akihide Ryo

## Abstract

SARS-CoV-2 neutralizing antibodies confer protective immunity against reinfection. We have developed a rapid test for screening SARS-CoV-2 neutralization antibodies using genome-free virus-like particles incorporated with a small luciferase peptide, HiBiT. Their entry into LgBiT-expressing target cells reconstitutes NanoLuc luciferase readily detected by a luminometer. This newly developed HiBiT-tagged Virus-like particle-based Neutralization Test (hiVNT) can readily quantify SARS-CoV-2 neutralizing antibodies within three hours with a high-throughput in a low biosafety setting. Moreover, the neutralizing activity obtained from hiVNT was highly consistent with that measured by the conventional neutralization test using authentic SARS-CoV-2. Furthermore, antibody responses to both viral spike and nucleocapsid proteins correlated with the neutralization activity assessed by hiVNT. Our newly-developed hiVNT could be instrumental to survey individuals for the presence of functional neutralizing antibody against SARS-CoV-2.

## Introduction

Due to the unavailability of any specific countermeasure, the constantly spreading COVID-19 pandemic could only be partially and temporarily slowed down by implementing regional lockdowns that force people to stay at home and prevent their movement. With the progression of the pandemic, a considerable subset of the population would have acquired post infection immunity and tests that reveal the post infection immune status of individuals are the need of the hour. A credible test which accurately identifies the protected can offer an immunity passport for the individual to be freed from the lockdown and resume routine activities without the fear of getting infected. At present, the semi-quantitative neutralization test (NT) and the quantitative plaque reduction neutralization test (PRNT) which identifies the presence of anti-SARS-CoV-2 neutralizing antibodies (nAbs) are the only foolproof methods available for this purpose. However, practical feasibility of these highly specific tests is weighed down by drawbacks such as; low throughput, long turnaround time (TAT) and the need for a specialized laboratory setup with biosafety level 3 (BSL3) facilities to handle the live viruses used in these tests ^1^.

To overcome these hurdles, pseudovirus-based NT ^2^, surrogate serodiagnostic tests (sVNT) ^3^, and single cell RNA sequencing ^1^ are being studied. However, a simple, convenient, rapid and high-throughput test capable of directly detecting nAbs with high specificity which could act as an ideal alternative to the neutralization assay is yet to be developed^4^. Virus-like particles (VLPs) are self-assembling, non-replicating, nonpathogenic entities of similar size and conformation as that of infectious virions. VLPs can be generated to possess the surface proteins of any kind of viruses on their membrane devoid of the viral nucleic acids. This genome-free feature of VLPs overrides the need for BSL3 facilities while handling, but has the drawback of difficulty in quantifying its cell entry and membrane fusion.

HiBiT is an 11 amino acid peptide tag that can be attached to any protein-of-interest. LgBiT, the counterpart of NanoLuc luciferase is complementary to HiBiT and binds to the latter to produce a highly active luciferase enzyme. HiBiT tagged proteins can be easily detected and quantified based on bioluminescence using the Nano-Glo assay system. HiBiT technology offers the advantages of high sensitivity, convenience of a single-reagent-addition step and short TAT of only a few minutes.

In this report, we have developed a HiBiT-VLP-based neutralization test (hiVNT) that can readily detect SARS-CoV-2 nAbs. The assay system utilizes VLPs bearing SARS-CoV-2 spike protein on their surfaces, incorporated with HiBiT-tagged HIV-1 capsid protein. Fusion of these VLPs could be readily monitored and quantified using VeroE6/TMPRSS2 cells stably expressing LgBiT as target cells. This system retains the specificity of neutralization assays while overcoming all the pitfalls, as hiVNT can be instrumental to measure protective immunity against viral invasion and evaluate vaccine efficacy by rapid, high-throughput quantification of nAb in a BSL2 laboratory.

## Results

To establish a rapid quantitative cell-based neutralization assay using luminescence-tagged VLPs, we inserted the HiBiT tag to the HIV-1 Gag-Pol protein, a minimal subunit to produce VLPs (Fig. 1A). After testing several prototypes, we found that HiBiT tag insertion at the C-terminal region of the capsid gene in Gag-Pol performed best. HiBiT tag can bind with high affinity to its complementary larger subunit LgBiT, and form luciferase (NanoLuc) ^5^. As expected, HiBiT-containing VLPs (hereafter designated as hiVLP-SARS2) could emit light upon addition of recombinant LgBiT and Furimazine substrate. As the capsid antigen carried the HiBiT tag, the luminescence intensity correlated with the amount of capsid antigen (Fig. 1B). By co-transfecting Gag-Pol-HiBiT and SARS-CoV-2 spike expression vectors, we generated hiVLP-SARS2 and confirmed their expression in cell and virus lysates (Fig. 1C).

**Figure 1.**
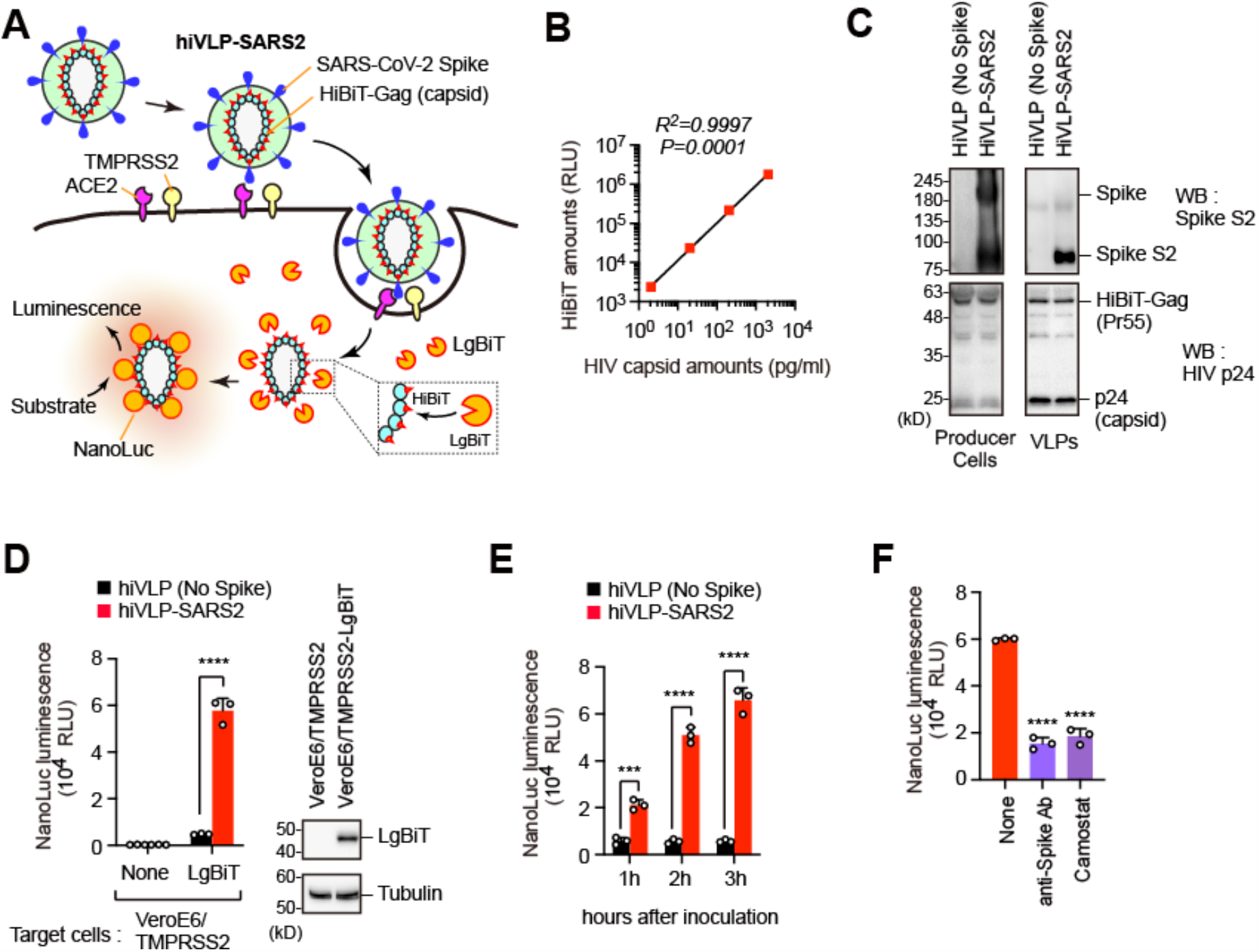
hiVLP-SARS2 system developed in this study. (**A**) Schematic representation of hiVLP-SARS2 system. (**B**) Incorporated HiBiT amounts in hiVLP-SARS2 correlate with p24 antigen of HIV-1. (**C**) Immunoblotting analysis of producer cells and hiVLP-SARS2. **(D, E)** Detection of cell entry of hiVLP-SARS2. VeroE6/TMPRSS2-LgBiT cells were inoculated with hiVLP-SARS2 for 3 hours (D) or 1-3 hours (E) after which the intracellular NanoLuc luminescence was measured. The expression of HaloTag-fused LgBiT in target cells were confirmed by immunoblotting analysis. \*\*\**P* < 0.001; \*\*\*\**P* < 0.0001. **(F)** VeroE6/TMPRSS2-LgBiT cells were inoculated with hiVLP-SARS2 for 3 hours in the presence of anti-Spike mAb (1:100, Sino Biological #40592-MM57) or Camostat (20 µM), after which the intracellular NanoLuc luminescence was measured. \*\*\*\**P* < 0.0001.

Since SARS-CoV-2 can infect VeroE6/TMPRSS2 cells with high efficiency ^6^, we next generated LgBiT-expressing VeroE6/TMPRSS2 cells. We noticed a robust increase in NanoLuc activity when the LgBiT-expressing VeroE6/TMPRSS2 cells were treated with hiVLP-SARS2 for 3 hours (Fig. 1D, E). We further demonstrated the drop in luminescence signals upon pretreatment with a TMPRSS2 inhibitor (Camostat mesylate) and also with anti-Spike neutralizing monoclonal antibody (Fig. 1F), suggesting that hiVLP-SARS2 enters the target cells through the interactions between viral spike and cellular receptors in a similar way as that of authentic SARS-CoV-2.

We next tested whether our newly developed hiVLP-SARS2 system could detect nAbs in the serum of COVID-19 patients. The neutralization assay was carried out in accordance with the following steps. VeroE6/TMPRSS2-LgBiT cells were seeded on 96-well white polystyrene plates at a density of 10^4^ cells/well one day prior to inoculation. Decomplemented sera derived from convalescent COVID-19 patients were mixed with hiVLP-SARS2 and the mixture was inoculated to VeroE6/TMPRSS2-LgBiT cells. Three hours later, the cells were washed with PBS and measured for their NanoLuc luminescence. We found that all the patients’ sera tested could block the entry of hiVLP-SARS2 in varying degrees (Fig. 2A), suggesting the presence of nAbs against SARS-CoV-2. We thus developed the hiVLP-SARS2-based Neutralization Test (hiVNT). Two convalescent sera and one healthy donor serum were tested by hiVNT and authentic SARS-CoV-2 NT only to show that all the samples gave similar results in both tests (Fig. 2B), suggesting that hiVNT performance is in concordance with the NT using authentic SARS-CoV-2.

**Figure 2.**
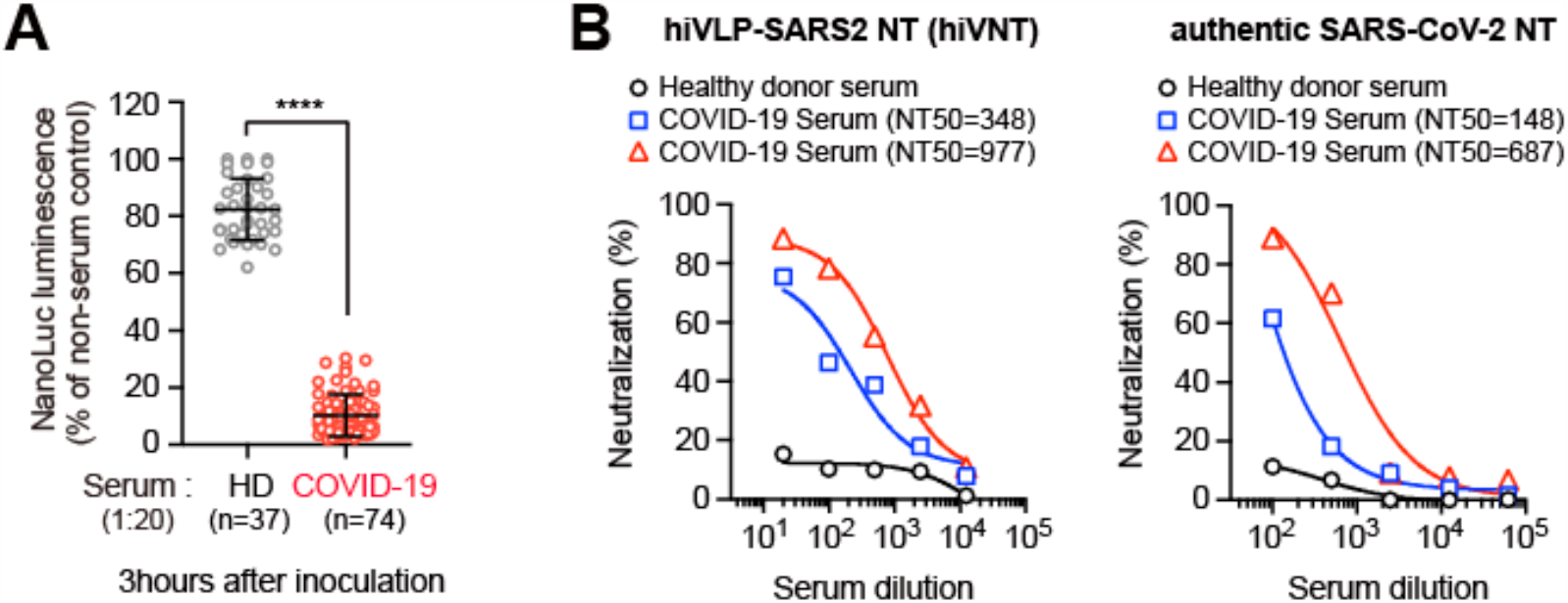
hiVLP-SARS2-based Neutralization Test (hiVNT). (**A**) NanoLuc luminescence of cells inoculated with hiVLP-SARS2 and either healthy donor (HD, N=37) or convalescent COVID-19 patient sera (N=74) for three hours. \*\*\*\**P* < 0.0001. (**B**) hiVNT is consistent with the NT using authentic SARS-CoV-2. Data of healthy donor and two representative patients are shown with NT titer (NT50).

Since surrogate antibody detection is being widely studied as an alternative to NT, we wanted to check the correlation of nAbs detected by hiVNT with antibody titers detected by ELISA. We matched the results of hiVNT against the antibody titers detected by ELISA in 74 COVID-19 positive sera. nAb levels were found to correlate with IgG antibodies against both the spike and the nucleocapsid proteins (Fig 3A), but not with IgM (Fig. 3B). Taken together, our novel hiVNT could be useful for rapid detection of nAbs in the sera of individuals recovered from COVID-19.

**Figure 3.**
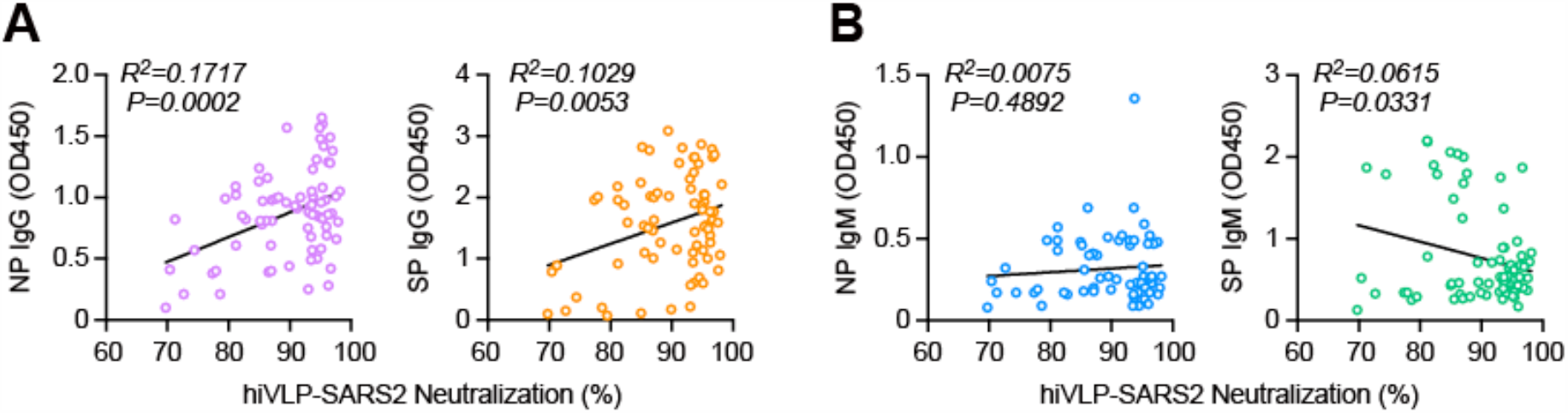
Correlation between serological test and neutralization activity of convalescent COVID-19 patient serum. (**A**) Correlation of neutralization activity with IgG against nucleocapsid (NP, left panel) or spike (SP, right panel). N=74 samples. (**B**) Correlation of neutralization activity with IgM against nucleocapsid (NP, left panel) or spike (SP, right panel). N=74 samples.

## Discussion

We have developed the hiVNT, a novel platform to rapidly detect and quantify SARS-CoV-2 nAbs using genome-free VLPs and HiBiT technology. The assay principle is similar to conventional neutralization assays and is based on viral entry and membrane fusion with measurement of NanoLuc luciferase activity to simplify the outcome. Considering the drawbacks of NT and PRNT, tests that may confer immunity passport to individuals are the need of the hour and different platforms are being exploited for this purpose, even though more detailed serological studies are needed.

Pseudovirus-based NTs are the popular alternatives to detect nAbs, as they overcome the need for BSL3 facility and have a high-throughput ^7^. These tests employ pseudoviruses with SARS-CoV-2 spike proteins for entry possessing a genome inserted with a reporter gene. The test depends on the expression of the reporter gene in the target cells which takes 24-48 hours to be detected. In comparison, our hiVNT does not involve gene expression and thus shortens the TAT to only around 3 hours. Single cell sequencing might have similar advantages ^1^ but the need for specialized and expensive equipment hinders its practical application.

Measurement of antiviral antibodies in the serum of convalescent COVID-19 patients is being studied for its potential to act as a surrogate marker to reflect the presence of SARS-CoV-2 nAbs ^3^. Our findings in a small sample size (n=74) reveal that IgG could serve as a better surrogate marker than IgM (Fig. 3). However, the use of serodiagnostics for nAb detection could have a few inherent drawbacks. Although all COVID-19 infections elicit a humoral immune response, the presence of antibodies does not reflect immunity. Also, mild COVID-19 infections elicit very low humoral response which might not be detected by serological tests ^4^. In this setting, surrogate antibody detection tests can produce erroneous results. Moreover, SARS-CoV-2 infects host cells through receptor binding domain (RBD) within S1 subunit in spike protein on the surface of viral particles that bind to host surface angiotensin converting enzyme-2 (ACE2) receptor. ELISA kits to detect anti-RBD antibodies use *in vitro* generated spike proteins and its derivatives to detect nAbs in serum. However, since spike proteins undergo glycosylation and oligomerization *in vivo* ^8,9^, the performance of *in vitro* generated spike proteins may vary depending on the manufacturing method, as well as their clinical significance. Also, nAbs that target other regions of spike protein to suppress the function of fusion peptide of S2 subunit may exist which could go undetected in S1 and RBD based detection ELISA ^10^.

In this study, we established the hiVNT; a simple, high-throughput assay system for the quantitative and rapid determination of SARS-CoV-2 nAbs in the sera of individuals after recovery from symptomatic or subclinical COVID-19 infection. The hiVNT system using VLPs, allows for BSL2-compliant testing and gives output in a short time. The concurrent use of the HiBiT system enhances the assay to give high-throughput quantitative results. We believe that the hiVNT can be instrumental in identifying individuals with protective immunity, epidemiological studies on population susceptibility, propagation modeling, assessing convalescent plasma used for therapy and vaccine evaluation; all these at a high-throughput and short TAT in a lower biosafety setting.

## Materials and Methods

### Ethics statement

This study was approved by Yokohama City University Certified Institutional Review Board (Reference No. B200200048, B160800009), and the protocols used in the study were approved by the ethics committee. Written informed consent was obtained from the patient or family/guardian.

### Cells and plasmids

HEK293 cells (ATCC #CRL-1573) and VeroE6/TMPRSS2 cells (JCRB #1819) were cultured in DMEM containing 10% FBS. VeroE6/TMPRSS2-LgBiT cells were generated by transfection of pCMV-HaloTag-LgBiT (Promega) with Neon Transfection System (Thermo) and selected with hygromycin (500 µg/ml). pHIV-GagPol-HiBiT was constructed by the insertion of HiBiT peptide sequence (VSGWRLFKKIS) into the C-terminal region of capsid (CA) gene of pHIV-GagPol, encoding codon-optimized HIV-1 Gag and Pol. SARS-CoV-2 Spike gene with replacement of the N-terminal signal peptide (residues 1–14) with CD5 signal sequence was synthesized and inserted into pcDNA6.2-FLAG vector. The spike gene lacks C-terminal ER retention signal (19 amino acids).

### Immunoblotting analysis

Samples in SDS loading buffer were loaded onto 10-20% polyacrylamide gels (Wako), electrophoresed, and blotted onto PVDF membranes (Merck). Membranes were probed with anti-SARS-CoV-2 Spike S2 (clone 1A9, GeneTex #632604) or anti-HIV p24 (clone 183-H12-5C, NIH AIDS Reagent Program #3537) antibodies and horseradish peroxidase-conjugated secondary antibodies (Cytiva). Detected proteins were visualized using LuminoGraph imaging system (ATTO).

### Production of hiVLP-SARS2

hiVLP-SARS2 was produced by transient transfection of HEK293 cells with pHIV-GagPol-HiBiT and pSARS2-Spike-FLAG at a ratio of 1:1. Culture supernatants containing VLPs were collected 48 hours after transfection and filtered through a 0.45 µm Millex-HV filter (Merck). Virion-incorporated HiBiT and p24 capsid antigens were measured by Nano-Glo HiBiT Lytic Detection System (Promega) and HIV-1 p24 antigen ELISA (Zepto Metrix), respectively.

### Neutralizing assay using hiVLP-SARS2 (hiVNT)

VeroE6/TMPRSS2-LgBiT cells seeded in 96-well plates were washed and inoculated with hiVLP-SARS2 stocks (50 µl) containing diluted serum (1:20 for Figs. 2A and 3, 1:20 to 1:12500 dilution for Fig. 2B). In some experiments, anti-spike neutralizing antibody (1:100, Sino Biological #40592-MM57) or Camostat mesilate (20 µM, Wako #039-17761) was added to the mixture instead of serum. At 3 hours after inoculation, cells were washed with PBS and treated with 100 µl of PBS containing DrkBiT peptide (1 µM) for 2 minutes. Cells were then added with 25 µl of 1x Nano-Glo Live Cell Substrate (Promega). Luciferase activity is measured with GloMax Discover System (Promega).

### Neutralizing assay using authentic SARS-CoV-2

SARS-CoV-2 (JPN/TY/WK-521) was obtained from National Institute of Infectious Diseases, Japan. For neutralizing assay, VeroE6/TMPRSS2 cells seeded in 96-well plates were washed and infected with 100 µl of medium containing SARS-CoV-2 (moi = 0.05) and five-fold serially diluted serum (1:100 to 1:62500 dilution). At 48 hours after infection, cells were washed and added with 40 µl of CellTiter-Glo Substrate (Promega). Cell viability is measured with GloMax Discover System (Promega).

### Neutralizing titer

The neutralizing titer (NT50) was calculated based on the half-maximal inhibitory concentrations determined by Prism 8.3.1 software (GraphPad).

The neutralization rate (%) was calculated as follows:

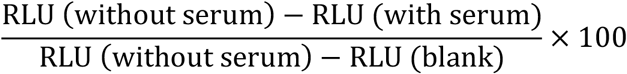

### ELISA

SARS-CoV-2 NP and SP recombinant protein were coated on 96-well plates (Nunc) overnight at 4°C. After blocking with 5% BSA in PBS for 1 hour at room temperature, diluted serum (1:100) were added to each well for 1 hour. Wells were then washed with PBS-T (PBS with 0.05% Tween-20) and added with HPR-conjugated goat anti-human IgM or IgG antibody, incubated for 1 hour at room temperature. Wells were washed with PBS-T and added 1-step TMB substrate (Thermo) and quenched the reaction with 0.5M H_2_SO_4_, followed by absorbance reading at 450 nm.

### Statistical analysis

All bar graphs present means and standard deviation. The statistical significance of differences between two groups was evaluated by two-tailed unpaired t-test in the Prism 8 software (GraphPad).

## Data Availability

All relevant data are available from the authors on request.

## Acknowledgement

We thank Chizu Suzuki, Kyoko Ohnishi, Kenji Yoshihara, and Kazuo Horikawa for their technical assistance.

## Funding

This study was supported by Rapid research and development Projects on COVID-19 of AMED (JP19fk0108110), and Life Innovation Platform Yokohama from Economic Affairs Bureau City of Yokohama to AR.

## Conflict of interests

The authors have no conflicts of interest directly relevant to the content of this article. YY is a current employee of Kanto Chemical Co. Inc.

## Author contributions

KM designed and performed the research, analyzed the data, and wrote the manuscript; SSJ analyzed the data, and wrote the manuscript; NO, SM, YY, M.Nishi, TM performed the research and analyzed the data; RS, M.Nishii, IT contributed reagents; HK, HH analyzed the data; AR directed the research, analyzed the data, and wrote the manuscript.

